# Differences in risk for SARS-CoV-2 infection among healthcare workers

**DOI:** 10.1101/2021.03.30.21254653

**Authors:** K. Miriam Elfström, Jonas Blomqvist, Peter Nilsson, Sophia Hober, Elisa Pin, Anna Månberg, Ville N. Pimenoff, Laila Sara Arroyo Mühr, Kalle Conneryd Lundgren, Joakim Dillner

## Abstract

Healthcare workers (HCWs) are a risk group for SARS-CoV-2 infection, but which healthcare work that conveys risk and to what extent such risk can be prevented is not clear. Starting on April 24^th^, 2020, all employees at work (n=15,300) at the Karolinska University Hospital, Stockholm, Sweden were invited and 92% consented to participate in a SARS-CoV-2 cohort study. Complete SARS-CoV-2 serology was available for n=12,928 employees and seroprevalences were analyzed by age, sex, profession, patient contact, and hospital department. Relative risks were estimated to examine the association between type of hospital department as a proxy for different working environment exposure and risk for seropositivity, adjusting for age, sex, sampling week, and profession. Wards that were primarily responsible for COVID-19 patients were at increased risk (adjusted OR 1.95 (95% CI 1.65-2.32) with the notable exception of the infectious diseases and intensive care units (adjusted OR 0.86 (95% CI 0.66-1.13)), that were not at increased risk despite being highly exposed. Several units with similar types of work varied greatly in seroprevalences. Among the professions examined, nurse assistants had the highest risk (adjusted OR 1.62 (95% CI 1.38-1.90)). Although healthcare workers, in particular nurse assistants, who attend to COVID-19 patients are a risk group for SARS-CoV-2 infection, several units caring for COVID-19 patients had no excess risk. Large variations in seroprevalences among similar units suggest that healthcare work-related risk of SARS-CoV-2 infection may be preventable.

## 1. Introduction

The first severe acute respiratory syndrome coronavirus 2 (SARS-CoV-2) outbreak emerged in late 2019 in China (1), and quickly developed into a global coronavirus disease (COVID-19) pandemic. Diagnostic surveillance of SARS-CoV-2 has reported more than 89.9 million SARS-CoV-2-infected and more than 1.9 million COVID-19 deaths. A major difficulty to prevent transmission of COVID-19 is that the pandemic is largely driven by asymptomatic and pre-symptomatic individuals, who are highly contagious (2, 3). Local, rapid COVID-19 outbreaks have been witnessed worldwide, particularly in elderly care homes (4, 5), nursing facilities (6) and among the healthcare workers (7-9).

Nosocomial infections notoriously not only endanger the patient but jeopardize healthcare personnel, and consequently restrain the capacity of a healthcare unit. Indeed, accumulating evidence indicates that healthcare workers (HCWs) are highly exposed to SARS-CoV-2 infections, and that a large fraction of the infections stem from asymptomatic individuals (10-13). Hence, it is of utmost importance to quickly and efficiently detect and manage healthcare-related COVID-19 exposures. To perform a reliable and large-scale assessment of nosocomial COVID-19 infections, we enrolled the staff of the Karolinska University Hospital in Stockholm, Sweden for SARS-CoV-2 antibody screening in April-June 2020, during the first COVID-19 outbreak. The risk of infection among the HCWs was estimated for each ward in the hospital and seroprevalences related to possible cofactors of infection, with particular focus on whether COVID-19 patients had been nursed at the wards.

## 2. Methods

### 2.1 Study population and setting

In response to the first wave of the COVID-19 pandemic, HCWs on duty at the Karolinska University Hospital (KUH) were invited to participate in a study that examined presence of antibodies to SARS-CoV-2 in serum. HCWs were recruited between April 23^rd^ and June 24^th^, 2020. Participants signed a written informed consent that included permission to link to the hospital administrative databases for information on professional role (physician, nurse/midwife, nurse assistant, or other), sick leave records and hospital unit assignment (s). The first COVID-19 patients were admitted to KUH in March 2020 and as the number of patients increased, hospital wards were re-purposed and healthcare workers were re-assigned to care for the large influx of COVID-19 patients.

The study was approved by the National Ethical Review Agency of Sweden (Decision number 2020-01620). Trial registration number: ClinicalTrials.gov NCT04411576.

### 2.2 Serological analyses

Whole blood was prepared as serum by centrifugation in serum-separating tubes at 2000 x g for 10 min. A heat treatment at 56 degrees C for 30 minutes was performed for virus inactivation and the samples were subsequently stored at −20 degrees C until further analysis.

SARS-CoV-2 IgG antibodies were analysed using three different virus protein variants, namely the trimeric full-length Spike protein (14) expressed in HEK cells, the Spike S1 domain expressed in CHO cells and the Nucleocapsid protein expressed in E. coli. The serum samples were processed as previously described (15) and analysed in a 384-plate format using a multiplex bead-based assay (Luminex corp.) with IgG detection towards all three viral antigens in parallel.

The performance of the serology assay was evaluated using 154 positive controls (defined as Covid-19 patients sampled at least 15 days after a positive PCR test) and 321 negative controls (defined as samples collected 2019 and earlier in the same region). The negative controls included 26 individuals with confirmed infections of other Coronaviruses than SARS-CoV-2. Based on these samples, the assay showed a 99.4% sensitivity and 99.1% specificity. Seropositivity was defined for each antigen as mean +6SD of 12 standardized negative control samples included in each analysis plate and for a sample to be assigned as IgG positive, a sample was required against at least two of the three included viral antigens. Captured IgG antibodies were detected by fluorescent anti-hIgG (Invitrogen, H10104) and reported as relative fluorescence intensity (AU).

### 2.3 Statistical analysis

For this study on risk of SARS-CoV-2 infection by type of healthcare work, participating healthcare workers were categorized based on their profession (physician, nurse/midwife, nurse assistant, or other), whether or not they had any patient contact, and their place of employment within the hospital. Hospital units were then categorized as intensive care units (ICU), COVID wards (meaning non-ICU units that cared for COVID-19 patients at some point during the study period) and “other” (including research, administrative and management units, that did not have routine contact with COVID-19 patients). Baseline data showing the distribution of HCW roles and places of employment within the hospital were estimated overall and by seroprevalence. Chi square tests and Cochran Armitage tests of trend were used to examine the association between covariates and seroprevalence. A multivariate logistic regression was run examining the association between type of hospital ward and serology positivity, adjusting for age, sex, sampling week, and professional role. Patient contact was not included in the model since it was strongly associated with the professional role of the HCWs. For the analysis of risk of SARS-CoV-2 infection associated with each hospital unit, HCWs with two or more affiliations listed were assigned to a ward-type using the following logic: HCWs were assumed to have contributed to COVID care, and then ICU or “other” units in that order given the need for additional support in the COVID wards. Hospital units with less than 50 participating HCWs were excluded from the unit-specific analysis, resulting in the removal of 44 units. The hospital units that cared for COVID-19 patients were noted and then serology positivity was reported as a prevalence with 95% confidence intervals. Using a case-cohort design, odds ratios were calculated comparing the odds of serology positivity in the unit to the odds of serology positivity in a sub-cohort consisting of all employees without any patient contact to estimate the excess risk of SARS-CoV-2 infection within each hospital unit.

The data were prepared and analyzed using SAS 9.4 Cary, NC, USA and a p-value of less than 0.05 was deemed significant.

## 3. Results

To assess risk of SARS-CoV-2 exposure by type of healthcare work, we enrolled 14,057 HCWs at work between April and June 2020 at Karolinska University Hospital (92% of all HCWs). We obtained valid serology results and formal employment status for 12,928 participants (Figure 1). Significant differences in Covid-19 seropositivity were observed by age, sampling (calendar) week, professional role of HCWs, and type of hospital unit (Table 1). A trend of decreased SARS-CoV-2 exposure infection with age from 16.4% (95% CI 14.6-18.3) among HCWs under the age of 30 to 8.6% (95% CI 7.4-9.9) among HCWs over the age of 60 was found (p-value for trend, <0.0001) (Table 1). Moreover, in the multivariate logistic regression model, a significant almost two-fold increased risk SARS-CoV-2 exposure was found for HCWs in hospital units caring for COVID-19 patients (adjusted OR 1.95 (95% CI 1.65-2.32)) as compared to units not caring for COVID-19 patients. Notably, ICU wards were not at an increased risk as compared to other wards despite intensive work with COVID patients (adjusted OR 0.86 (95% CI 0.66-1.13)). Differences in risk for seropositivity were also seen by profession and sampling week. Nurse assistants had the highest risk for seropositivity (adjusted OR 1.62 (95% CI 1.38-1.90)), followed by nurses and midwives (adjusted OR 1.28 (95% CI 1.11-1.49)) as compared to a reference group of other healthcare personnel. Physicians did not have a significantly increased risk. By calendar time, the highest risk for SARS-CoV-2 seropositivity was observed in the last weeks of sampling in the study (calendar weeks 21+) as compared to the first weeks (calendar weeks 17-18) (adjusted OR 1.45 (95% CI 1.24-1.71)) (Table 2).

**Figure 1.**
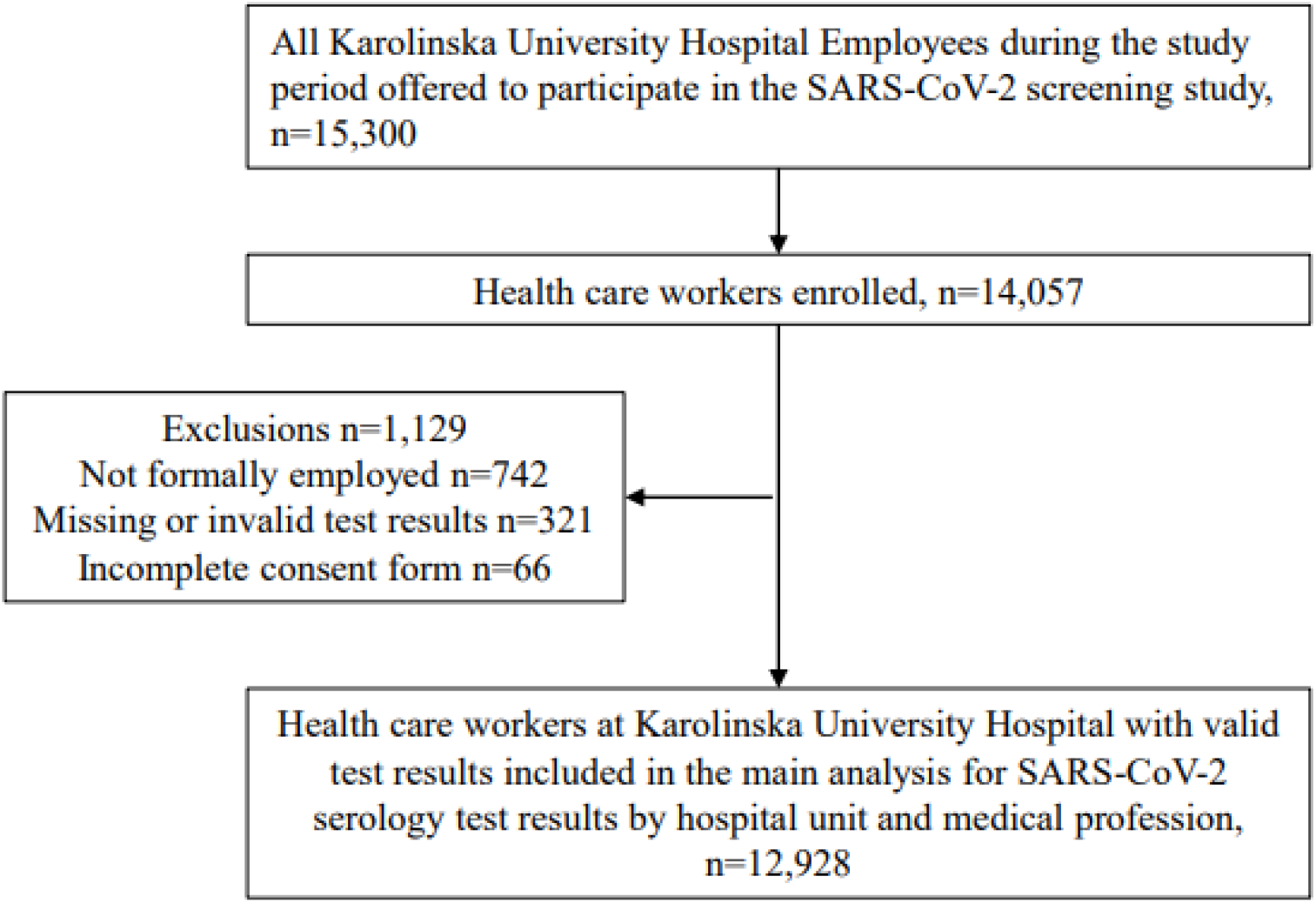
STROBE flowchart of study participants.

**Table 1.** Distribution of background covariates among 12,928 employees of the Karolinska University Hospital.

|  |  |  |  |
| --- | --- | --- | --- |
|  | n (%) | Serology positive<br>n (% , 95% CI) | p-value |
| Age |  |  |  |
| ≤29 | 1,522 (11.8) | 249 (16.4, 14.6-18.3) | <0.0001* |
| 30-39 | 3,172 (24.5) | 382 (12.0, 11.0-13.2) |  |
| 40-49 | 3,238 (25.1) | 370 (11.4, 10.4-12.6) |  |
| 50-59 | 3,066 (23.7) | 313 (10.2, 9.2-11.3) |  |
| ≥60 | 1,930 (14.9) | 166 (8.6, 7.4-9.9) |  |
| Sex |  |  |  |
| Female | 10,203 (78.9) | 1,170 (11.5, 10.9-12.1) | 0.8945** |
| Male | 2,725 (21.1) | 310 (11.4, 10.2-12.6) |  |
| Sampling week |  |  |  |
| Week 17-18 | 6,801 (52.6) | 699 (10.3, 9.6-11.0) | <0.0001* |
| Week 19-20 | 4,274 (33.1) | 539 (12.6, 11.7-13.6) |  |
| Week 21+ | 1,853 (14.3) | 242 (13.1, 11.6-14.7) |  |
| Profession |  |  |  |
| Other personnel | 4,043 (31.3) | 385 (9.5, 8.7-10.5) | <0.0001** |
| Nurse/midwife | 4,027 (31.2) | 486 (12.1, 11.1-13.1) |  |
| Nurse assistant | 2,532 (19.6) | 389 (15.4, 14.0-16.8) |  |
| Physician | 2,326 (18.0) | 220 (9.5, 8.3-10.7) |  |
| Patient contact |  |  |  |
| No patient contact | 3,671 (28.4) | 356 (9.7, 8.8-10.7) | <0.0001** |
| Patient contact | 9,257 (71.6) | 1,124 (12.1, 11.5-12.8) |  |
| Type of hospital unit |  |  |  |
| Other | 11,259 (87.1) | 1,192 (10.6, 10.0-11.2) | <0.0001** |
| COVID-19 ward | 1,031 (8.0) | 225 (21.8, 19.4-24.5) |  |
| Intensive care unit (ICU) | 638 (4.9) | 63 (9.9, 7.8-12.4) |  |

\*Cochran Armitage Trend Test
\*\*Chi square test

**Table 2.**
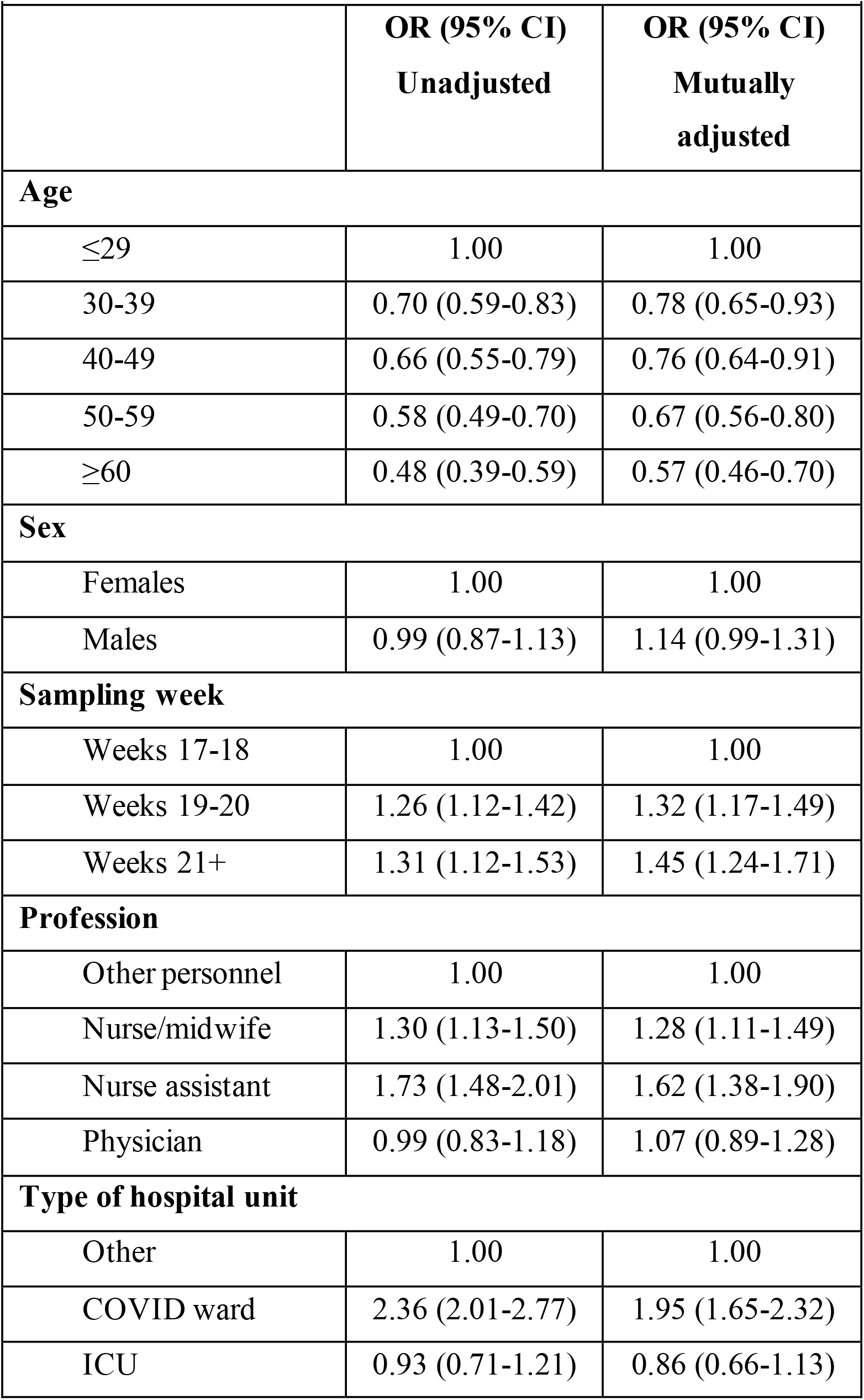
Association between type of hospital ward and serology positivity, adjusted for co-factors.

|  |  |  |
| --- | --- | --- |
|  | **OR (95% CI)**<br>**Unadjusted** | **OR (95% CI)**<br>**Mutually adjusted** |
| **Age** |  |  |
| ≤29 | 1.00 | 1.00 |
| 30-39 | 0.70 (0.59-0.83) | 0.78 (0.65-0.93) |
| 40-49 | 0.66 (0.55-0.79) | 0.76 (0.64-0.91) |
| 50-59 | 0.58 (0.49-0.70) | 0.67 (0.56-0.80) |
| ≥60 | 0.48 (0.39-0.59) | 0.57 (0.46-0.70) |
| **Sex** |  |  |
| Females | 1.00 | 1.00 |
| Males | 0.99 (0.87-1.13) | 1.14 (0.99-1.31) |
| **Sampling week** |  |  |
| Weeks 17-18 | 1.00 | 1.00 |
| Weeks 19-20 | 1.26 (1.12-1.42) | 1.32 (1.17-1.49) |
| Weeks 21+ | 1.31 (1.12-1.53) | 1.45 (1.24-1.71) |
| **Profession** |  |  |
| Other personnel | 1.00 | 1.00 |
| Nurse/midwife | 1.30 (1.13-1.50) | 1.28 (1.11-1.49) |
| Nurse assistant | 1.73 (1.48-2.01) | 1.62 (1.38-1.90) |
| Physician | 0.99 (0.83-1.18) | 1.07 (0.89-1.28) |
| **Type of hospital unit** |  |  |
| Other | 1.00 | 1.00 |
| COVID ward | 2.36 (2.01-2.77) | 1.95 (1.65-2.32) |
| ICU | 0.93 (0.71-1.21) | 0.86 (0.66-1.13) |

To comprehensively explore the risk of SARS-CoV-2 exposure for HCWs in different hospital settings, we compared the odds of seropositivity in each hospital unit to the odds of seropositivity in a reference group of hospital personnel without patient contact (Table 3). HCWs in six out of ten COVID wards had more than two-fold increased risk of SARS-CoV-2 exposure compared to HCWs without patient contact. The highest, almost three-fold increased risk for SARS-CoV-2 exposure was observed for HCWs in units that had been repurposed to nurse COVID-19 patients but that would typically care for patients with non-infectious diseases (e.g. neurology, geriatrics, and cardiology). There were 5/55 wards who had not nursed COVID-19 patients but still had a similar, nearly two-fold increased risk of SARS-CoV-2 exposure.

**Table 3.** Serology positivity by hospital unit, odds of serology positivity for each hospital unit compared to odds among participants with no patient contact.

| **Hospital unit** | **n serology positive/total # part. in unit** | **% (95% CI)** | **OR (95% CI) Unadjusted** |
| --- | --- | --- | --- |
| §Pelvic cancer | 52/361 | 14.4 (11.2-18.4) | 1.52 (1.11-2.08) |
| §Upper abdominal cancers | 23/198 | 11.6 (7.9-16.8) | 1.18 (0.76-1.86) |
| §Breast cancers, endocrine tumors and sarcomas | 31/141 | 22.0 (15.9-29.5) | 2.54 (1.68-3.85) |
| §ENT, hearing and balance | 23/231 | 10.0 (6.7-14.5) | 1.00 (0.64-1.56) |
| §Cardiology, ward | 53/213 | 24.9 (19.6-31.1) | 2.99 (2.15-4.16) |
| §Geriatrics, ward | 46/185 | 24.9 (19.2-31.6) | 2.99 (2.10-4.25) |
| §Inflammation and infectious diseases, ward | 45/343 | 13.1 (10.0-17.1) | 1.36 (0.98-1.90) |
| §Neurology, ward | 54/207 | 26.1 (20.6-32.5) | 3.19 (2.29-4.43) |
| §Emergency care, ward 1 | 50/249 | 20.1 (15.6-25.5) | 2.27 (1.63-3.16) |
| §Emergency care, ward 2 | 82/405 | 20.3 (16.6-24.4) | 2.29 (1.75-3.00) |
| §ICU & thorax surgery | 59/760 | 7.8 (6.1-9.9) | 0.76 (0.57-1.01) |
| §Pediatric ICU | 38/317 | 12.0 (8.9-16.0) | 1.23 (0.86-1.76) |
| §Neonatology | 35/367 | 9.5 (6.9-13.0) | 0.95 (0.66-1.37) |
| R&D management, Division for research | 6/93 | 6.5 (3.0-13.4) | 0.62 (0.27-1.44) |
| R&D management, Division for education | 6/86 | 7.0 (3.2-14.4) | 0.68 (0.29-1.56) |
| ICU, Resident physicians | 1/65 | 1.5 (0.3-8.2) | 0.14 (0.02-1.02) |
| Development and administration | 3/63 | 4.8 (1.6-13.1) | 0.45 (0.14-1.45) |
| Human resources | 6/101 | 5.9 (2.8-12.4) | 0.57 (0.25-1.31) |
| Staffing administration | 16/111 | 14.4 (9.1-22.1) | 1.52 (0.88-2.61) |
| Clinical nutrition | 7/50 | 14.0 (7.0-27.2) | 1.47 (0.66-3.29) |
| Social work | 5/85 | 5.9 (2.5-13.0) | 0.56 (0.23-1.40) |
| Occupational therapy and physiotherapy | 17/195 | 8.7 (5.5-13.5) | 0.86 (0.52-1.44) |
| Emergency care | 42/303 | 13.9 (10.4-18.2) | 1.45 (1.03-2.05) |
| Neuroradiology | 4/96 | 4.2 (1.6-10.2) | 0.39 (0.14-1.08) |
| Medical radiation and nuclear medicine | 11/119 | 9.2 (5.2-15.8) | 0.92 (0.49-1.73) |
| Clinical physiology | 7/69 | 10.1 (5.0-19.5) | 1.02 (0.46-2.25) |
| Radiology 1 | 17/166 | 10.2 (6.5-15.8) | 1.03 (0.62-1.72) |
| Radiology 2 | 9/96 | 9.4 (5.0-16.9) | 0.93 (0.47-1.87) |
| Perioperative medicine 1 | 40/396 | 10.1 (7.5-13.5) | 1.01 (0.72-1.43) |
| Perioperative medicine 2 | 25/148 | 16.9 (11.7-23.8) | 1.84 (1.18-2.86) |
| Newborn screening lab | 4/70 | 5.7 (2.2-13.8) | 0.55 (0.20-1.51) |
| Pathology and cytology | 40/400 | 10.0 (7.4-13.3) | 1.00 (0.71-1.42) |
| Clinical genetics | 5/79 | 6.3 (2.7-14.0) | 0.61 (0.24-1.52) |
| Clinical pharmacology | 6/137 | 4.4 (2.0-9.2) | 0.41 (0.18-0.94) |
| Clinical immunology and transfusion medicine | 36/292 | 12.3 (9.0-16.6) | 1.11 (0.77-1.60) |
| Clinical chemistry | 22/320 | 6.9 (4.6-10.2) | 0.67 (0.43-1.04) |
| Clinical microbiology | 18/266 | 6.8 (4.3-10.4) | 0.66 (0.40-1.07) |
| Point-of-care laboratory network | 24/237 | 10.1 (6.9-14.6) | 1.02 (0.66-1.57) |
| Cardiology | 22/183 | 12.0 (8.1-17.5) | 1.23 (0.78-1.95) |
| Infectious diseases | 4/61 | 6.6 (2.6-15.7) | 0.63 (0.23-1.76) |
| Gastric, skin. and rheumatic disorders | 11/165 | 6.7 (3.8-11.5) | 0.65 (0.35-1.20) |
| Endocrinology | 5/83 | 6.0 (2.6-13.3) | 0.58 (0.23-1.44) |
| Nephrology | 9/159 | 5.7 (3.0-10.4) | 0.54 (0.27-1.07) |
| Gynecology and reproductive medicine | 17/95 | 17.9 (11.5-26.8) | 1.97 (1.15-3.37) |
| Neurology | 3/65 | 4.6 (1.6-12.7) | 0.44 (0.14-1.40) |
| Radiation | 5/86 | 5.8 (2.5-12.9) | 0.56 (0.22-1.39) |
| Hematology | 14/169 | 8.3 (5.0-13.4) | 0.82 (0.47-1.43) |
| Head & neck, lung and skin cancers | 14/204 | 6.9 (4.1-11.2) | 0.67 (0.38-1.16) |
| Transplantation medicine | 6/75 | 8.0 (3.7-16.4) | 0.79 (0.34-1.82) |
| Plastic surgery and craniofacial diseases | 12/74 | 16.2 (9.5-26.2) | 1.75 (0.93-3.28) |
| Pediatric surgery and medicine, ward | 25/202 | 12.4 (8.5-17.6) | 1.28 (0.83-1.97) |
| Pediatric emergency care, ward | 24/185 | 13.0 (8.9-18.6) | 1.35 (0.86-2.10) |
| Pregnancy and deliveries, ward | 52/400 | 13.0 (10.1-16.7) | 1.35 (0.99-1.84) |
| Pediatric surgery and medicine, ward | 16/95 | 16.8 (10.6-25.6) | 1.83 (1.06-3.17) |
| Pediatric orthopedics, ward | 15/130 | 11.5 (7.1-18.2) | 1.18 (0.68-2.04) |
| Neurosurgery, ward | 8/66 | 12.1 (6.3-22.1) | 1.25 (0.59-2.63) |
| Management, Imaging & Functional diagnostics div. | 12/123 | 9.8 (5.7-16.3) | 0.98 (0.53-1.79) |
| Management, Hospital Laboratory div. | 12/71 | 16.9 (9.9-27.3) | 1.84 (0.98-3.45) |
| Management, ICU div. | 20/132 | 15.2 (10.0-22.3) | 1.62 (0.99-2.63) |
| Management, Geriatrics div. | 6/58 | 10.3 (4.8-20.8) | 1.04 (0.44-2.44) |
| Management, Inflammatory & Infectious diseases | 5/76 | 6.6 (2.8-14.5) | 0.64 (0.26-1.59) |
| Management, Pediatrics div. | 34/277 | 12.3 (8.9-16.7) | 1.26 (0.87-1.84) |
| Management, Gynecology & Obstetrics div. | 6/99 | 6.1 (2.8-12.6) | 0.58 (0.25-1.34) |
| Management, Cardiology & Neurology div. | 13/70 | 18.6 (11.2-29.2) | 2.06 (1.12-3.80) |
| Management, Cancer div. | 4/106 | 3.8 (1.5-9.3) | 0.35 (0.13-0.97) |
| Management, emergency care and surgery div. | 8/100 | 8.0 (4.1-15.0) | 0.79 (0.38-1.63) |
| Support and informatics | 47/280 | 16.8 (12.9-21.6) | 1.82 (1.31-2.54) |
| Innovation and development | 15/158 | 9.5 (5.8-15.1) | 0.95 (0.55-1.63) |

§Wards that cared for COVID-19 patients

## 4. Discussion

### 4.1 Statement of the main findings

We found that wards that were caring for COVID-19 patients were at increased risk (adjusted OR 1.95 (95% CI 1.65-2.32) with the notable exception of the infectious diseases and intensive care units (adjusted OR 0.86 (95% CI 0.66-1.13)), that were not at increased risk despite being highly exposed. We also report that units with similar types of work may vary greatly in seroprevalences and that the profession associated with highest risk was nurse assistant (adjusted OR 1.62 (95% CI 1.38-1.90)).

### 4.2 Strengths and considerations

Our study is quite large, and we enrolled the majority of the employees (>92% (12,928/15,300)) of the Karolinska University Hospital suggesting that the data are robust. The study exploited the fact that the linkable hospital administrative systems contained pre-collected and exact information on e.g. patient contact, place of work, profession, and the repurposing of hospital units for COVID-19 care. HCWs can, of course, be exposed to SARS-CoV-2 outside the hospital (community transmission). However, the fact that seropositivities clustered in wards re-purposed for COVID-19 care implies that transmission from patients to HCWs must have occurred at least once in such wards. Once introduced, it is of course possible that the HCWs working in the same unit can transmit infections to each other. The fact that some non-COVID-19 wards also had high seroprevalences could be compatible with outbreaks that had been introduced to the units by community transmission.

### 4.3 Comparison with others

A seroprevalence survey among HCWs (n=46,117) at a New York City-based health system found a 13.7% seropositivity rate overall and that 9% of asymptomatic HCWs were also seropositive (10). Among employees at 22 elderly care centers in Sweden, 23.0% (231/1,005) were seropositive and 46.5% of the seropositive employees did not report symptoms (16). Another Swedish cohort study of seroprevalence among HCWs at another acute care hospital (n=2,149) that was performed during the same time period and used exactly the same serology method found a seropositivity of 19.1% and an association with patient contact (OR 2.9, 95% CI 1.9–4.5) (11).

## 5. Conclusion

We find that SARS-CoV-2 transmission was not random, and instead, we found clear clustering of SARS-CoV-2 exposure among HCWs in specific hospital wards. In agreement with previous reports, our results clearly suggest that nosocomial transmission of SARS-CoV-2 is common, in particular among nurse assistants in wards re-purposed to care for COVID-19 patients. Importantly, however, we also show that examples that HCWs in hospital wards prepared for infectious diseases control and caring for high amount of COVID patients did not have increased risk for SARS-CoV-2 exposure. The large variability among units also suggests that with appropriate preventative measures healthcare work-related risks of SARS-CoV-2 infection may be significantly reduced. In severe outbreak situations, HCWs are likely to be more exposed. As the seroprevalences among HCWs in different units varied widely, it is possible that stringent workplace preventive measures may reduce this risk.

## Data Availability

Data can be made available upon request to Dr. Joakim Dillner.

## CRediT authorship contribution statement

K. Miriam Elfström (Data curation, Formal analysis, Writing – original draft); Jonas Blomqvist (Data curation, Formal analysis, Writing – review and editing); Peter Nilsson (Funding acquisition, Investigation, Project administration, Resources, Supervision, Writing – review and editing); Sophia Hober (Investigation, Resources, Writing – review and editing); Elisa Pin (Investigation, Writing – review and editing); Anna Månberg (Investigation, Writing – review and editing); Ville N. Pimenoff (Writing – original draft); Laila Sara Arroyo Mühr (Investigation, Writing – review and editing); Kalle Conneryd Lundgren (Conceptualization, Funding acquisition, Resources, Writing – review and editing); and Joakim Dillner (Conceptualization, Funding acquisition, Project administration, Resources, Supervision, Writing – original draft).

## Acknowledgments

Support was obtained from the Karolinska University Hospital, the County Council of Stockholm, Erling-Persson family foundation, KTH Royal Institute of Technology, Creades, and SciLifeLab. The funding agencies have had no role in the design, execution or interpretation of the study or in the decision to submit for publication.

We would like to thank Suyesh Amatya, Helena Andersson, Shaghayegh Bayati, Emine Eken, Pedram Farsi, Yasmin Hussein, Roxana Merino Martinez, Sara Mravinacova, Björn Pfeifer, Ulla Rudsander, Sadaf Sakina Hassan, Ronald Sjöberg, Balazs Szakos, Hanna Tegel, and Emel Yilmaz for excellent technical assistance.

